# Analysis of Social Combinations of Coronavirus Vaccination: Evidence from a Conjoint Analysis

**DOI:** 10.1101/2021.06.13.21258654

**Authors:** Hanako Ohmura

**Affiliations:** School of Policy Studies, Kwansei Gakuin University. 2-1, Gakuen, Sanda-shi, Hyougo-ken, Japan, 6691337

**Keywords:** vaccination, wait-and-see strategy for vaccination, herd immunity, familiar entity, conjoint analysis

## Abstract

Using a conjoint analysis based on cases in Japan, this study attempts to identify a preferable social strategic combination of “Who is vaccinated, who is not, who waits.” The analysis shows that the most desirable choice is a “wait-and-see” strategy, allowing for a risk assessment of side effects. We also find that subjects who recalled blood relatives as their familiar entities tend to prefer a “wait-and-see” strategy for themselves and their blood relatives.

## 1 Introduction

With the spread of the Coronavirus (COVID-19), consequent deaths, and after-effects in many countries, the best hope for mass immunization is vaccine development and administration. However, in society, individual intent to receive vaccination remains insufficient. Representative studies show that the percentage of individuals willing to be vaccinated is 53.6% (undecided: 14.4%, unwilling: 32%) in the United States (Daly and Robinson, 2021) and 73.9% (undecided: 18.9%, unwilling: 7.2%) in European countries (Neumann-Böhme et al., 2020), which are the main suppliers or initiators of vaccinations.^1^

Several studies have been conducted based on surveys of intended vaccination. Studies examining vaccination intent in more detail aim to determine the vaccine properties that individuals prefer, immunization protocols in place, and willingness of individuals to be vaccinated according to these situations (for example McPhedran and Toombs, 2021; Motta, 2021; Kreps et al., 2020). These studies adopted discrete choice experiments (McPhedran and Toombs, 2021) or conjoint analyses (Motta, 2021) to identify the vaccine property preferences of individuals, to detect the requirements for increased vaccine uptake. In addition to the research on these vaccines as “one product,” using the above methods, it may be necessary to consider the influence of vaccination status in society on the intention of an individual to be vaccinated. One problem that previous studies do not address is the need to examine the status of strategic interactions, namely, preferences of who in a society should be inoculated and who should not, about the individual and their loved ones.^2^

While many individuals will value scientific evidence and find it desirable to vaccinate themselves altruistically, and vaccinate the entire community, the most desirable combination of vaccinations among themselves, familiar individuals, and society overall, can occur in various ways.^3^ In addition, some individuals may think strategically, wishing to “receive the vaccine after confirming the mid-/long-term adverse reactions,” rather than a binary choice of simply wishing to receive it or not. Considering this nuanced alternative also allows us to capture certain egoistic (individually rational) attitudes, such as “society as a whole should be vaccinated early, and I should be vaccinated after confirming the side effects and the effect on the variant.” A strategic conflict between altruism and egoism is likely to arise in a society that provides vaccines, requiring a framework to explain its consequences.

Similar to previous studies, this study uses the choice-based conjoint analysis for vaccina-tion, to identify the types of strategic vaccine combinations most desirable in society, considering Japan.

The expectation was that the Japanese case would provide meaningful insights. Japan is a latecomer to the vaccine market, only starting to vaccinate healthcare professionals on February 18, 2021. Vaccinations have begun with less uncertainty regarding side effects than in other countries. In addition, as Yoda and Katsuyama (2021) report, of the 1,100 online survey participants in Japan, 65.7% were willing to be vaccinated, 22.0% were not sure, and 12.3% were not willing to be vaccinated. Therefore, Japan has a high proportion of individuals adopting a “waitand-see policy” due to the US and European countries starting vaccinations earlier. Japan’s case has important implications for ascertaining the social impact of the vaccination program, when many individuals have access to information about its side effects, and with a low level of uncertainty about the vaccine.

The next section describes the design of the conjoint analysis. Section 3 summarizes the results of this analysis. Based on the results, Section 4 discusses the implications of this study.

## 2 Empirical Strategy

### 2.1 Study subjects and period

This study aims to identify vaccine combinations by introducing conjoint analysis into an online survey. It conducted two online survey experiments, guiding survey panels from the Yahoo Crowd Sourcing Inc. (YCS) (March 14–16, 2021), and Lucid Holdings LLc. (March 26–28, 2021). Lucid had 1,024 subjects and 27 questions, while YCS had 2,975 subjects and 43 questions. Subjects were based on quotas allocated according to their demographic composition.^4^

In addition, considering the inoculation schedule, we considered this survey period appro-priate for verifying Japan’s vaccination status. The vaccination of healthcare workers began on February 18, 2021, and we conducted our studies after about one month. Around that time, it was reported that a woman died of a cerebrovascular disease after vaccination. Also at about the same time, at a meeting of the vaccine study group held by the Ministry of Health, Labour and Welfare, several cases of adverse reactions caused by anaphylactic shock were reported. Considering these series of events, we conducted our survey approximately when Japanese citizens decided upon vaccination; that is, when more information about vaccination *per se* and its side effects was available.^5^

### 2.2 Design of conjoint analysis

The contrivance in this conjoint analysis is as follows. First, to prevent an attribute of the conjoint from becoming too complicated, the pattern of attributes was constituted into three main bodies of “myself,” “familial presence,” and “society as a whole.” Subjects are asked to select one of the following, “familial presence, as a *reference group*: a family member who is older than the subject (e.g. parents), a family member who is younger than the subject (e.g. children), friend, colleague, neighbor, spouse, and significant other/partner.^6^ The subjects were instructed to face the conjoint while recalling a selected alternative as a familiar presence.

Second, the level for each attribute should not be a binary choice between vaccination and non-vaccination, as a third option should be incorporated, namely, “vaccinate later.” This alternative allows us to capture certain egoistic attitudes, such as, “society as a whole should be vaccinated early, and we should be vaccinated after confirming the occurrence of side effects.” If the effect of *a watcher* is greater than that of a simple desire for “I will vaccinate,” the acquisition of herd immunity by vaccine is not necessarily an optimistic scenario.

According to the above conjoint analysis settings, we set our design of conjoint as in Table 1 and Figure 1, and Figure 2shows an example of the conjoint screen.

**Table 1:** Proportion of willingness to vaccinate

| Attributes | Values |  |  |
| --- | --- | --- | --- |
| Myself | Vaccinate | Do not vaccinate now, vaccinate later | Not vaccinate |
| Familiar presence | Vaccinate | Do not vaccinate now, vaccinate later | Not vaccinate |
| Society as a whole | Vaccinate | Do not vaccinate now, vaccinate later | Not vaccinate |

**Figure 1:**
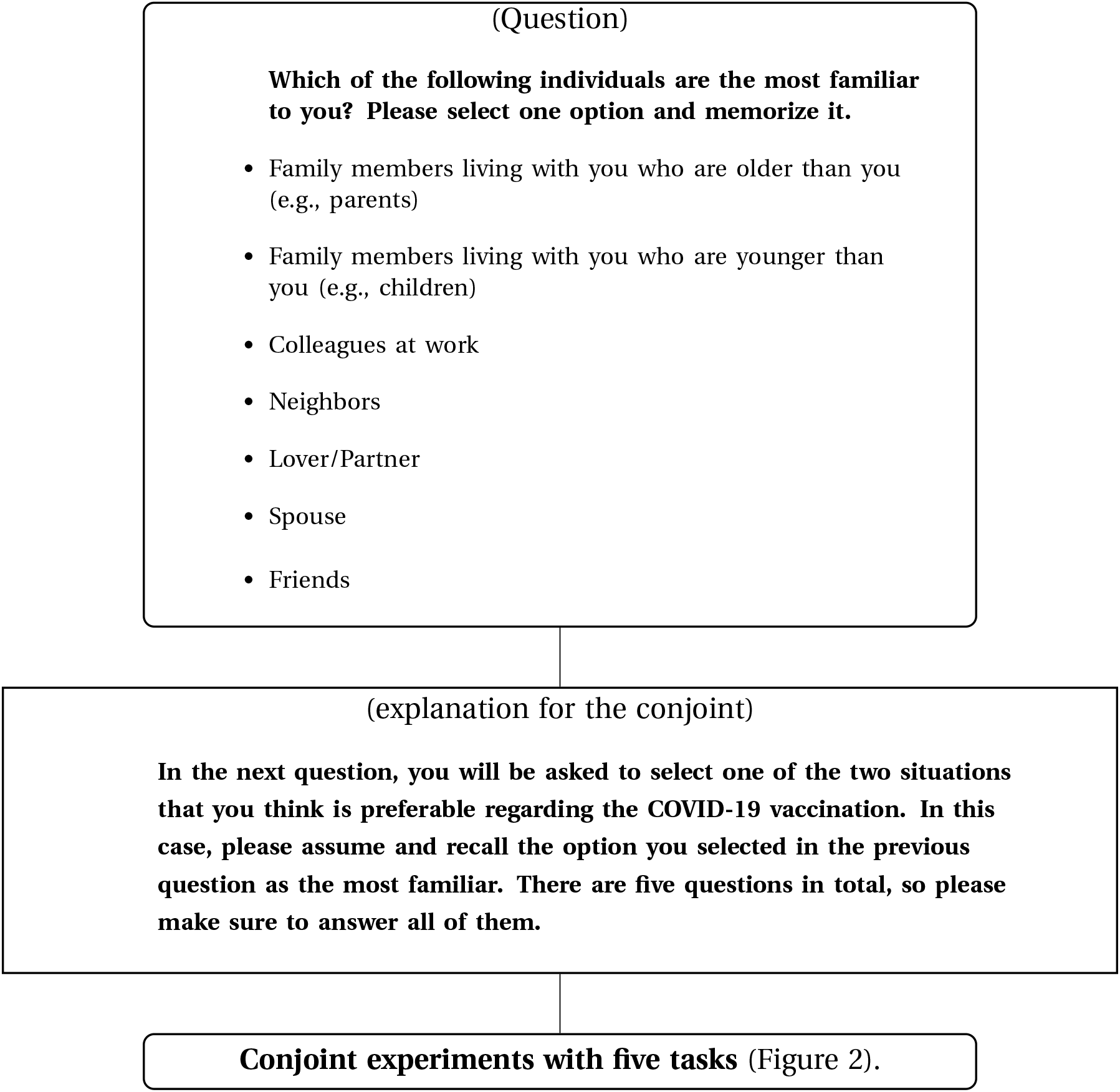
Conjoint experiment flow

**Figure 2:**
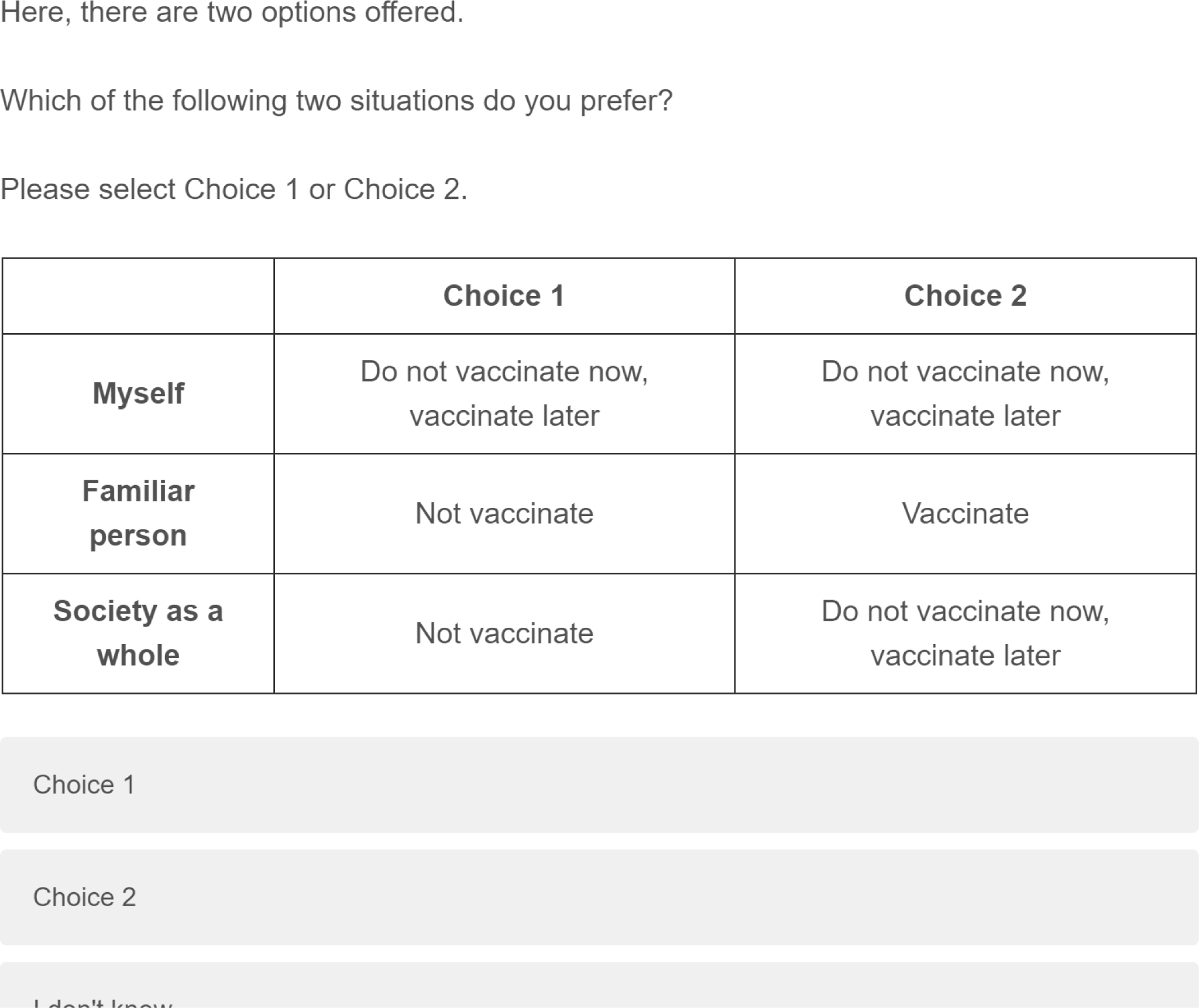
Display example of conjoint analysis. *Note*: Subjects were randomly displayed five tasks.

We performed conjoint analysis using the Conjoint Survey Design Tool and introducing the conjoint program into Qualtrics, according to the procedure by Hainmueller et al. (2014). Using the method in Hainmueller et al. (2014), it is possible to measure the effect of the concerned attribute X under all other attributes, as the average marginal component effect (AMCE), even if the effect of attribute X of interest is heterogeneous with regard to the distribution of other attributes. Considering the examples in this study, Hainmueller (2014) enables us to measure the effect of the “myself” intention to vaccinate, based on the overall effects across other attributes: society as a whole and familiar entity.

## 3 Results

First, Table 2 provides the simple descriptive statistics on the intention to take the vaccine.^7^ According to the YCS results, the most common response was, “vaccinate later,” followed by, “receive vaccination,” and finally, “not receive vaccination.” Lucid’s result implies that vaccinations exceeded “wait-and-see” vaccinations; however, many people in Japan still have a “wait-and-see” strategy. We may have these results partly because we conducted the survey immediately after the reporting of specific information on adverse reactions, and it is likely that more citizens in Japan prefer a wait-and-see strategy than in other countries. A major difference from the results of Yoda and Katsuyama (2021) is expected in the reporting of adverse reactions and the stabilization of infections during the investigation period.^8^

**Table 2:** Proportion of willingness to vaccinate

|  | Vaccination | Wait-and-see | No vaccination |
| --- | --- | --- | --- |
| YCS | 0.418 | 0.449 | 0.133 |
| Lucid | 0.462 | 0.401 | 0.137 |

In addition, the majority of the respondents who answered “vaccination later” or “no vaccination,” said they wanted to check the side effects of the vaccine; as predicted, the percentage exceeded 50% in all surveys (Figure 3). The next is an altruistic reason: “there are people who should get vaccinated before me.” By contrast, the third most common reason was “once herd immunity is established, it is not necessary to inoculate yourself,” which was marked as a more strategic and egoistic intent.

**Figure 3:**
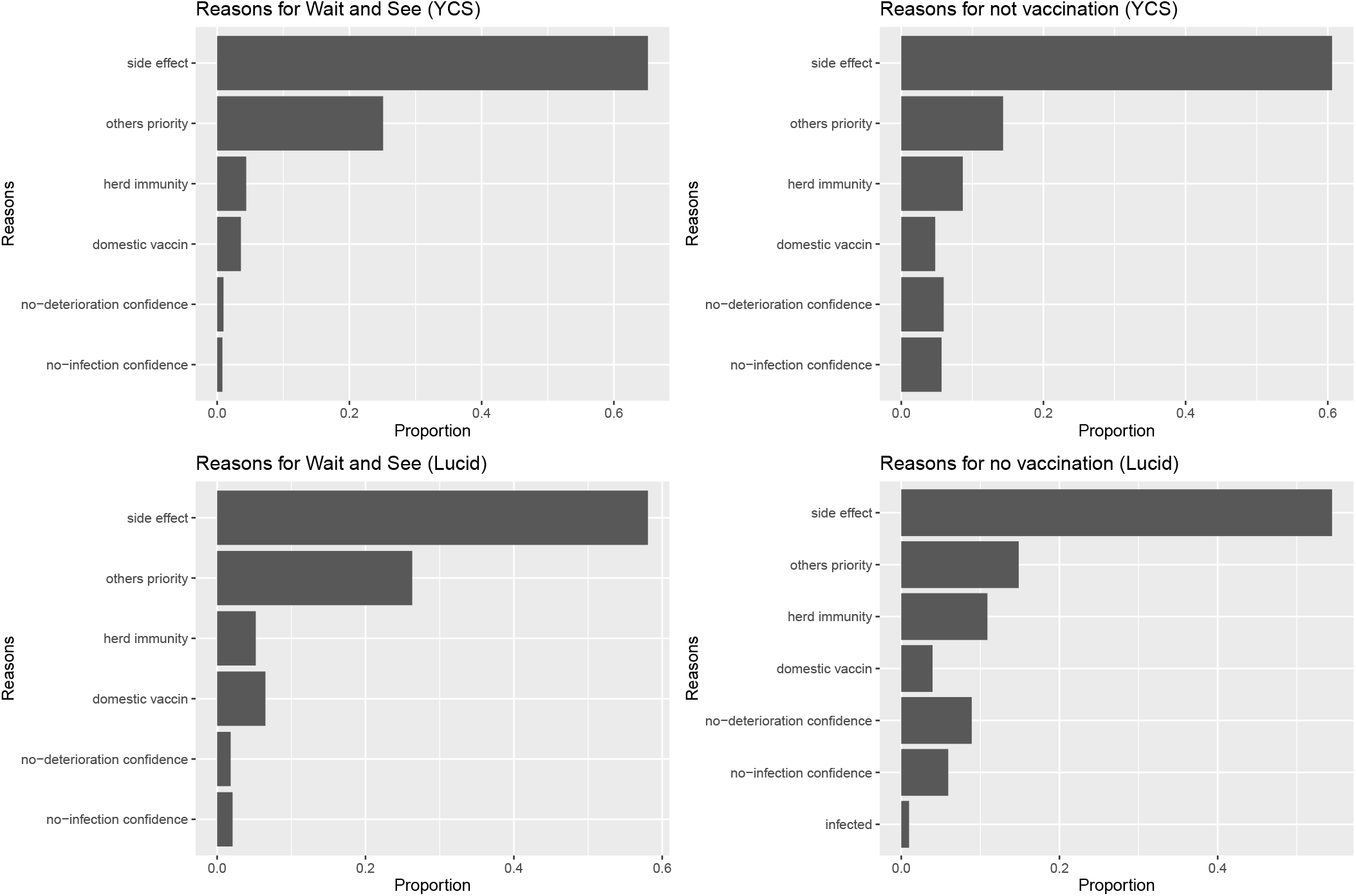
Reasons for not vaccinating and watchers. *Note*:Abbreviations: side effects=I want to see if there are any adverse reactions to the vaccine. Others priority=The product of the desired vaccine manufacturer is not available in Japan.Herd immunity=If others are inoculated first and herd immunity is established, there is no need to inoculate myself as soon as possible. No-deterioration confidence=Even if I am infected, it is unlikely that I will become seriously ill. No-infection confidence=I will not be infected. Infected=I have already been infected with COVID-19.

We then examined the results of a pooled conjoint analysis of all subjects. Figure 4 provides stronger support for descriptive analysis, revealing that “wait-and-see” is the preferred strategy for self and personal existence (baseline: not vaccination). It is difficult to find significant differences between the overall results of Lucid’s panel in the AMCEs because of the small sample size. However, there are significant differences among AMCEs: vaccination, watchers, and non-vaccination, especially, in the case of familiar entities. Furthermore, for society as a whole, the results show no noticeable difference in the effects of vaccination among watchers. By contrast, for society in general, the results show no difference between static and active inoculation. Both results for familiar entities and society suggest that the *altruism* of “withholding vaccination for the time being” seems to work not for society in general, but only for others who are particularly close. The results of this study showed no support for altruism, which is meant to contribute to herd immunity by actively inoculating oneself. More importantly, it is clear that a “wait-and-see” strategy is preferred for oneself and those close to oneself, whereas the watcher strategy for society as a whole is not.

**Figure 4:**
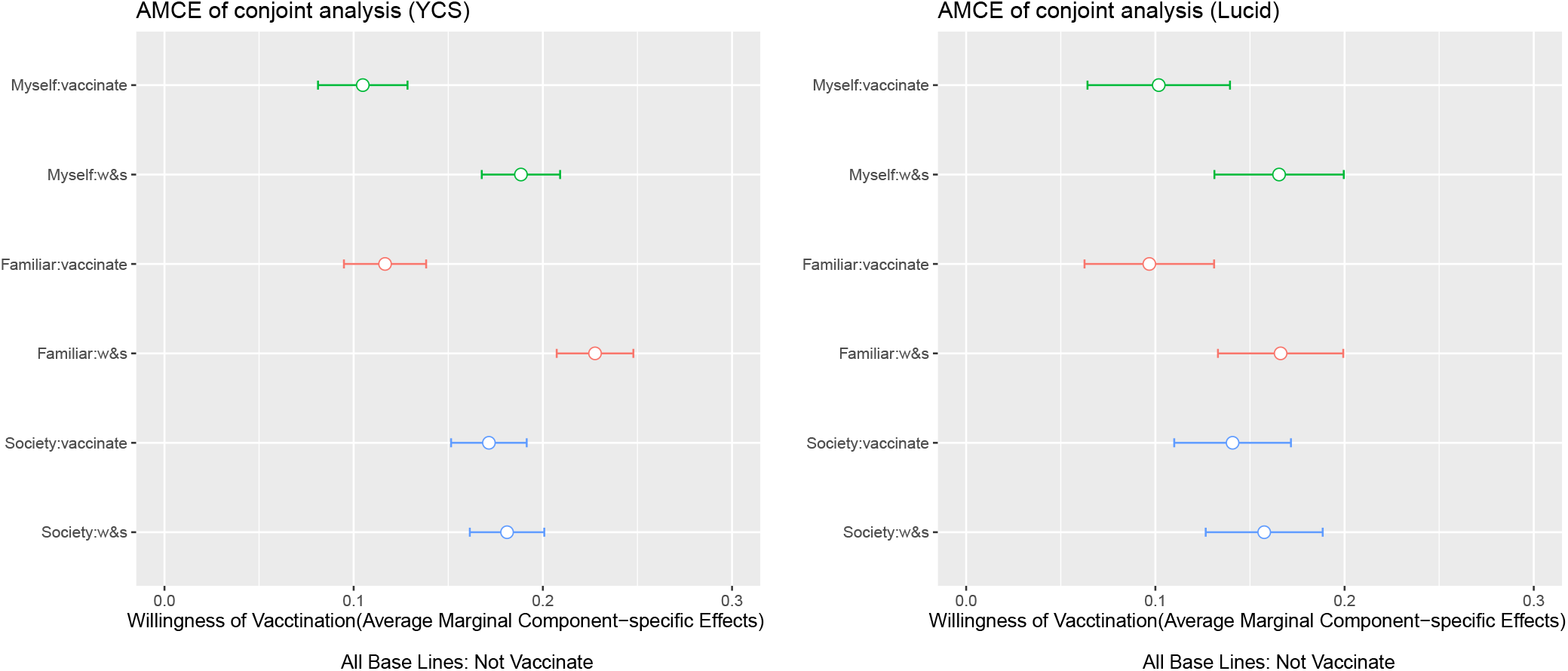
Average marginal component effect (AMCE) results. *Note*: This plot shows estimates of the effects of the randomly assigned vaccination attribute values on the probability of being preferred by Japanese subjects. The estimates are based on the benchmark OLS model with clustered standard errors, and the bars represent 99% confidence intervals. The points denote the attribute value, which is the reference category for each attribute.

Next, we examined the conjoint results relating to the familiar entities. As Figure 5 shows, the most common choices in descending order were as follows: spouse, family members living together who were older and younger than the subjects. This order was the same in both studies. Clearly, close family was the most frequent choice. In this context, Figure 6 shows the results of conjoint sorted according to who is selected as a familiar entity, and its baseline is set as not to be inoculated. Strikingly, those who select younger family members are less likely to prefer vaccinations for familiar entities, and are more likely to hope that younger members are watchers. Furthermore, even the group that selected elderly living family members as the highest risk group at the time of infection, did not prefer vaccination among those close to them (especially, in Lucid’s case), and a “wait-and-see” strategy played a central role.

**Figure 5:**
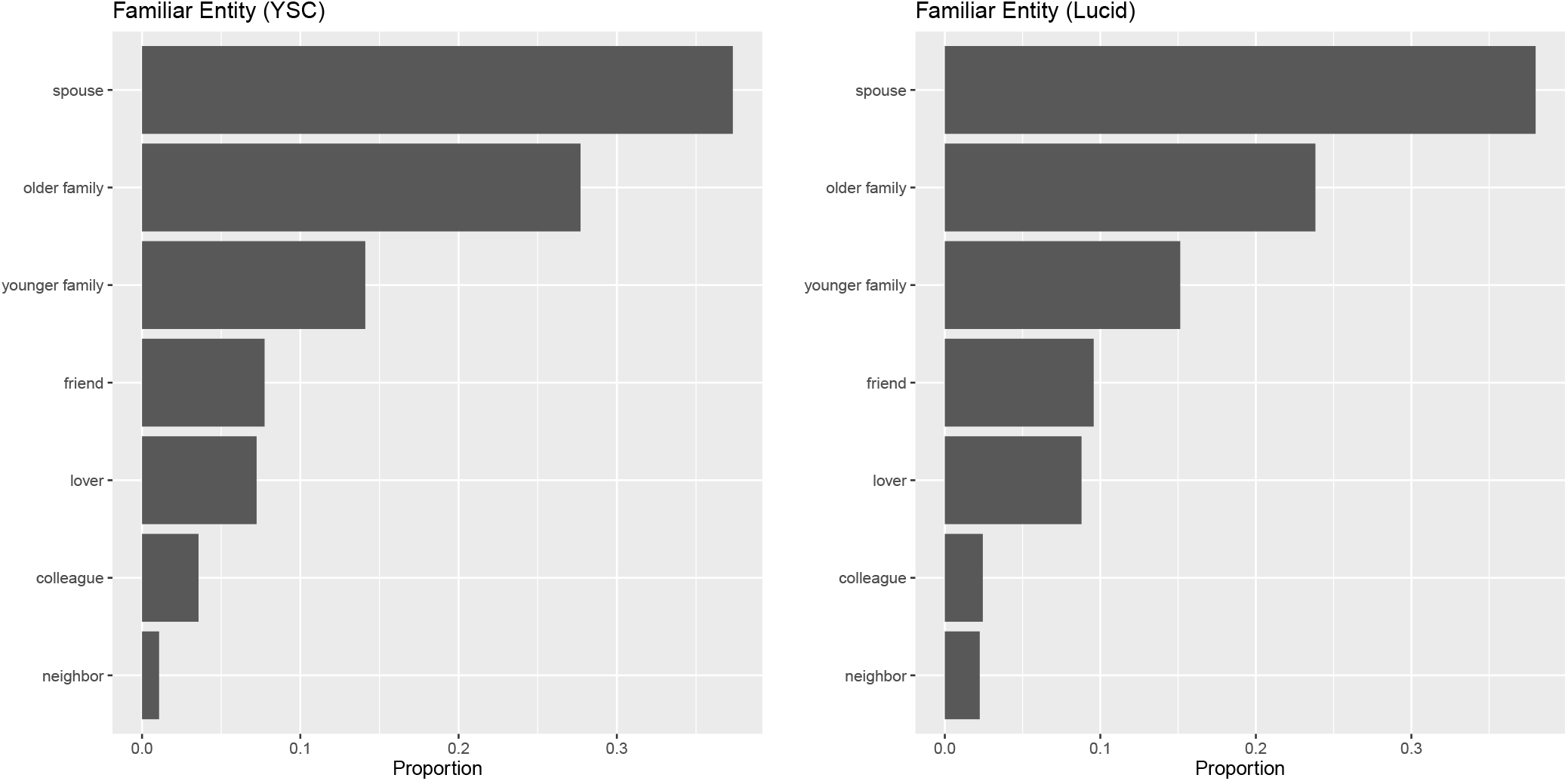
Selection of familiar entities

**Figure 6:**
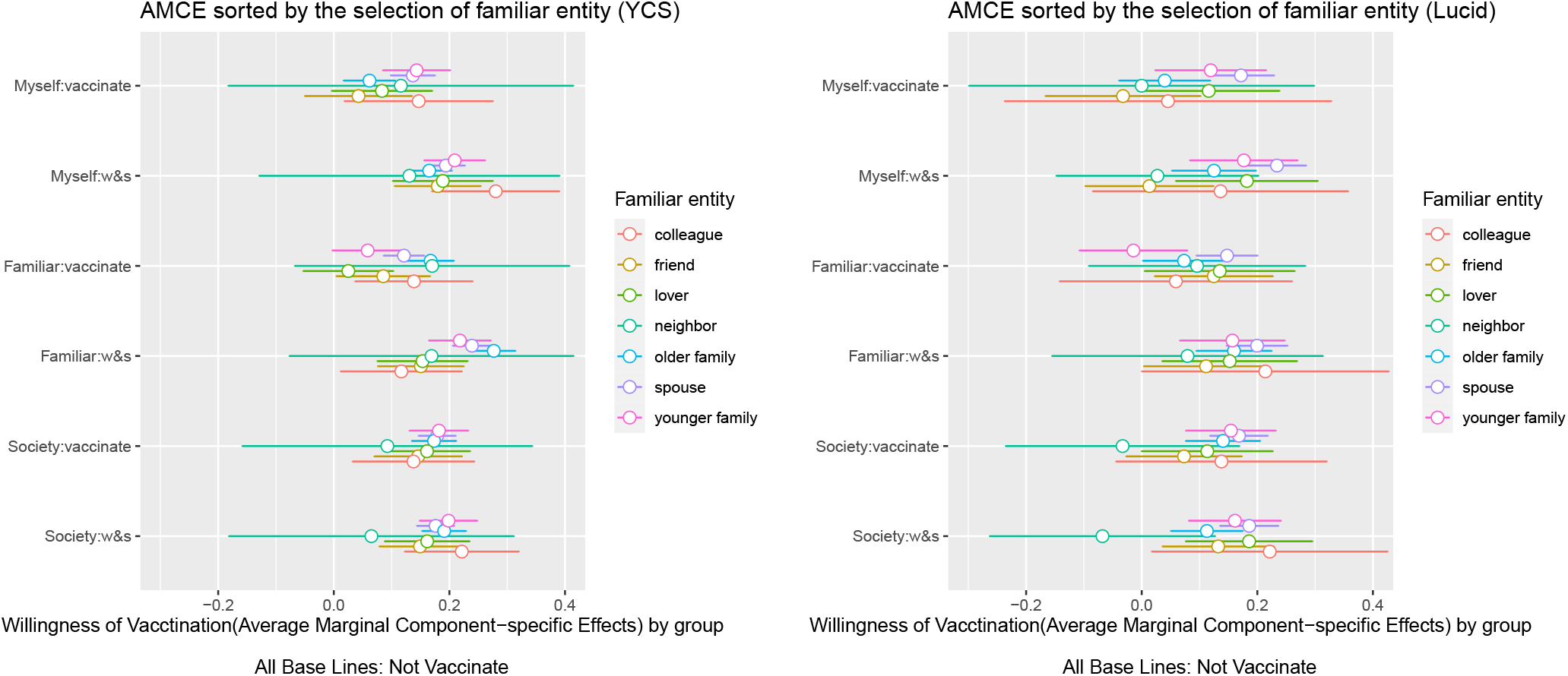
Average marginal component effect (AMCE) results by group. *Note*: This plot shows estimates of the effects of the randomly assigned vaccination attribute values on the probability of being preferred by Japanese subjects. Coefficient plots are sorted by groups, as per familial presence recalled by subjects. The estimates are based on the benchmark OLS model with clustered standard errors, and the bars represent 99% confidence intervals. The points denote the attribute value, which is the reference category for each attribute.

## 4 Conclusions

In Japan, with fewer COVID-19 deaths than in other countries, intention to receive the vaccination is immature, with individuals adopting a “wait-and-see” strategy. A conjoint analysis, further delving into intentions, showed a strong preference for a “wait-and-see strategy,” not only for themselves but also for those they were familiar with. This suggests that altruism does not lead to herd immunity through active voluntary vaccinations, but reduces the side effects of vaccination, the objective being to protect self or familiars. In addition, this study reveals that the preference for a familiar to adopt a “wait-and-see” strategy was stronger in those who recall blood relations as familiar entities. Our analysis indicates that the most desirable social combinations of vaccination are as follows: society as a whole may or may not be vaccinated early, but my loved ones and I can wait long enough to be vaccinated.

These results suggest that it is difficult to establish herd immunity by increasing the number of vaccinated individuals at an early stage. To suppress opportunistic strategies and cultivate intentions for mass vaccination, it is imperative to inform the public fully that the risk of infection is higher than the risk of vaccine side effects.

## Supporting information

https://www.dropbox.com/s/i4qm0o89ehnzgd9/supplymentary_covid19_202106.pdf?dl=0

## Data Availability

All data is available upon requests.

## Footnotes

1 Daly’s (2020) survey was conducted twice in April and October 2020. The sample size was 7547. It reported that the number of individuals wanting to receive the vaccine decreased from 71% in April to 53.6% in October. Neumann-Böhme et al. (2020) collected 1,000 samples each from Denmark, France, Germany, Italy, Portugal, the Netherlands, and the UK and conducted an additional survey of 500 samples from Lombardy in April 2020.

2 A survey of healthcare workers asking their children whether they would like to be vaccinated includes (Gold-man et al., 2020).

3 One study found that altruistic vignettes were more effective in enhancing vaccination intent Rieger (2020).

4 Supplemental Materials contain descriptive statistics on respondent varying variables and demographic composition compared to census.

5 Regarding ethics, the study was undertaken by the Kwansei Gakuin University Committee for Regulations for Behavioral Research with Human Participants. In accordance with the Committee’s recommendations, subjects were informed at the beginning of the survey that they may refuse to be presented with sensitive information about the novel coronavirus and may leave the investigation at any time. They were also informed at the debriefing that if they felt uncomfortable with the information they received, they could opt not to send their responses. The compensation for the survey was set at 20 Yahoo points for YCS and 3.8 USD for Lucid.

6 For the criterion for distinguishing familiar individuals, we refer to the reference group set up in a study of relative income with the hypothetical choice experiment by Clark and Senik (2010) and Yamada and Sato (2016).

7 The results of the Test of Equal or Given Proportions indicate no difference between the two surveys in the three options (vaccination: *p =* 0.0449, wait-and-see: *p =* 0.1431, no vaccination: *p =* 0.4325).

8 However, we assumed that the vaccination intentions in this study are much higher at this point (as of June 2021) for several reasons. First, the spread of infection and the collapse of medical care caused by the British variant since April. Second, the government is greatly accelerating vaccination ahead of the Tokyo Olympics in July. Third, vaccination is being expanded to non-elderly populations through large-scale vaccination using companies and universities. On the other hand, it has also been reported that vaccination has not necessarily progressed contrary to previous predictions, and a proportion of the population may still be holding the “wait-and-see strategy.

