## Supplementary material for "Analysis of Social Combinations of Coronavirus Vaccination: Evidence from a Conjoint Analysis": https://www.dropbox.com/s/i4qm0o89ehnzgd9/supplymentary_covid19_202106.pdf?dl=0

Evidence from Conjoint analysis

June 9, 2021

### 1 Descriptive Statistics and Demographic Composition

Table 1: Descriptive statistics of demography (Yahoo Crowd Sourcing)

| | n | mean | sd | median | trimmed | mad | min | max | range | skew | kurtosis |
|---|---|---|---|---|---|---|---|---|---|---|---|
| age | 2927 | 46.22 | 11.25 | 46.00 | 46.29 | 10.38 | 4.00 | 86.00 | 82.00 | -0.05 | 0.01 |
| gender (female=1) | 2934 | 0.41 | 0.60 | 0.00 | 0.35 | 0.00 | 0.00 | 4.00 | 4.00 | 2.19 | 9.45 |
| education (4year=1) | 2896 | 0.57 | 0.50 | 1.00 | 0.59 | 0.00 | 0.00 | 1.00 | 1.00 | -0.28 | -1.92 |
| income | 2934 | 602.77 | 432.23 | 526.00 | 552.71 | 335.07 | 0.00 | 3000.00 | 3000.00 | 2.03 | 7.40 |

Table 2: Descriptive statistics of demography (Lucid)

| | n | mean | sd | median | trimmed | mad | min | max | range | skew | kurtosis |
|---|---|---|---|---|---|---|---|---|---|---|---|
| age | 994 | 44.59 | 16.15 | 44.00 | 44.19 | 19.27 | 8.00 | 86.00 | 78.00 | 0.16 | -0.85 |
| gender (female=1) | 1024 | 1.53 | 0.55 | 2.00 | 1.51 | 1.48 | 1.00 | 4.00 | 3.00 | 0.61 | 0.78 |
| education (4year=1) | 1010 | 0.53 | 0.50 | 1.00 | 0.54 | 0.00 | 0.00 | 1.00 | 1.00 | -0.14 | -1.98 |
| income | 1024 | 869.09 | 654.91 | 684.50 | 777.85 | 493.71 | 0.00 | 3000.00 | 3000.00 | 1.32 | 1.54 |

Table 3: Comparison between survey demographic and national census (Yahoo Crowd Sourcing)

| | | Census | YCS | $t$-test results |
|---|---|---|---|---|
| Age | | | | |
| | 20-25 | 0.07 | 0.11 | $t = 0$ |
| | 25-30 | 0.07 | 0.08 | $p$-value = 1 |
| | 30-35 | 0.08 | 0.11 | |
| | 35-40 | 0.09 | 0.10 | |
| | 40-45 | 0.10 | 0.11 | |
| | 45-50 | 0.11 | 0.11 | |
| | 50-55 | 0.10 | 0.10 | |
| | 55-60 | 0.09 | 0.12 | |
| | 60-65 | 0.09 | 0.07 | |
| | 65-70 | 0.09 | 0.07 | |
| | 70-75 | 0.11 | 0.04 | |
| Gender | | | | |
| | Female | 0.51 | 0.38 | $t = 0.0003$ |
| | Male | 0.49 | 0.62 | $p$-value = 0.9998 |
| Education | | | | |
| | Elementary | 0.19 | 0.00 | $t = 0$ |
| | High Schoole | 0.46 | 0.24 | $p$-value = 1 |
| | Two-year college | 0.15 | 0.19 | |
| | Four-year college and graduate school | 0.20 | 0.57 | |
| Income | | | | |
| | 0-100 | 0.07 | 0.02 | $t = 0.136$ |
| | 100-200 | 0.13 | 0.07 | $p$-value = 0.893 |
| | 200-300 | 0.14 | 0.12 | |
| | 300-400 | 0.13 | 0.12 | |
| | 400-500 | 0.10 | 0.12 | |
| | 500-600 | 0.09 | 0.14 | |
| | 600-700 | 0.07 | 0.08 | |
| | 700-800 | 0.06 | 0.08 | |
| | 800-900 | 0.05 | 0.08 | |
| | 900-1000 | 0.04 | 0.04 | |
| | 1000-1100 | 0.05 | 0.03 | |
| | 1100-1400 | 0.03 | 0.05 | |
| | 1400-1600 | 0.05 | 0.02 | |
| | 1600-1800 | 0.01 | 0.01 | |
| | 1800-2000 | 0.00 | 0.00 | |
| | 2000- | 0.01 | 0.02 | |

Table 4: Comparison between survey demographic and national census (Lucid)

| | | Census | Lucid | $t$-test results |
|---|---|---|---|---|
| Age | | | | |
| | 20-25 | 0.07 | 0.02 | $t = 0$ |
| | 25-30 | 0.07 | 0.04 | $p$-value = 1 |
| | 30-35 | 0.08 | 0.08 | |
| | 35-40 | 0.09 | 0.12 | |
| | 40-45 | 0.10 | 0.16 | |
| | 45-50 | 0.11 | 0.21 | |
| | 50-55 | 0.10 | 0.15 | |
| | 55-60 | 0.09 | 0.11 | |
| | 60-65 | 0.09 | 0.07 | |
| | 65-70 | 0.09 | 0.04 | |
| | 70-75 | 0.11 | 0.02 | |
| Gender | | | | |
| | Female | 0.51 | 0.50 | $t = 0.003$ |
| | Male | 0.49 | 0.50 | $p$-value = 0.998 |
| Education | | | | |
| | Elementary | 0.19 | 0.02 | $t = 0$ |
| | High School | 0.46 | 0.26 | $p$-value = 1 |
| | Two-year college | 0.15 | 0.19 | |
| | Four-year college and graduate school | 0.20 | 0.53 | |
| Income | | | | |
| | 0-100 | 0.07 | 0.02 | $t = 0.168$ |
| | 100-200 | 0.13 | 0.04 | $p$-value = 0.868 |
| | 200-300 | 0.14 | 0.09 | |
| | 300-400 | 0.13 | 0.09 | |
| | 400-500 | 0.10 | 0.09 | |
| | 500-600 | 0.09 | 0.11 | |
| | 600-700 | 0.07 | 0.06 | |
| | 700-800 | 0.06 | 0.08 | |
| | 800-900 | 0.05 | 0.05 | |
| | 900-1000 | 0.04 | 0.06 | |
| | 1000-1100 | 0.05 | 0.04 | |
| | 1100-1400 | 0.03 | 0.09 | |
| | 1400-1600 | 0.05 | 0.05 | |
| | 1600-1800 | 0.01 | 0.03 | |
| | 1800-2000 | 0.004 | 0.03 | |
| | 2000- | 0.01 | 0.08 | |
